# Pediatric brain tumor classification using deep learning on MR-images from the children’s brain tumor network

**DOI:** 10.1101/2023.05.12.23289829

**Authors:** Tamara Bianchessi, Iulian Emil Tampu, Ida Blystad, Peter Lundberg, Per Nyman, Anders Eklund, Neda Haj-Hosseini

**Author notes:** corresponding author: Iulian Emil Tampu. Authors contributed equally (Tamara Bianchessi, Iulian Emil Tampu).

## Abstract

**Background and purpose:** Brain tumors are among the leading causes of cancer deaths in children. Initial diagnosis based on MR images can be a challenging task for radiologists, depending on the tumor type and location. Deep learning methods could support the diagnosis by predicting the tumor type.

**Materials and methods:** A subset (181 subjects) of the data from “Children’s Brain Tumor Network” (CBTN) was used, including infratentorial and supratentorial tumors, with the main tumor types being low-grade astrocytomas, ependymomas, and medulloblastomas. T1w-Gd, T2-w, and ADC MR sequences were used separately. Classification was performed on 2D MR images using four different off-the-shelf deep learning models and a custom-designed shallow network all pre-trained on adult MR images. Joint fusion was implemented to combine image and age data, and tumor type prediction was computed volume-wise. Matthew’s correlation coefficient (MCC), accuracy, and F1 scores were used to assess the models’ performance. Model explainability, using gradient-weighted class activation mapping (Grad-CAM), was implemented and the network’s attention on the tumor region was quantified.

**Results:** The shallow custom network resulted in the highest classification performance when trained on T2-w or ADC MR images fused with age information, when considering infratentorial tumors only (MCC: 0.71 for ADC and 0.64 for T2-w), and both infra- and supratentorial tumors (MCC: 0.70 for ADC and 0.57 for T2-w).

**Conclusion:** Classification of pediatric brain tumors on MR images could be accomplished using deep learning, and the fusion of age information improved model performance.

## Introduction

Tumors in the central nervous system are the second most common type of cancer in children and young adults up to the age of 19, with an estimated age-standardized rate (in 100,000 population) of 1.2 for incidence and 0.59 for mortality worldwide^1^, where brain tumors account for about 57% of the total causes of cancer deaths in this population^2^. Pediatric brain tumors (PBT) can be grouped concerning the location relative to the tentorium, as infratentorial or supratentorial. Tumors in the infratentorial brain region (posterior fossa) are more common in pediatric patients; however, the frequency varies depending on age^3–5^. Brain tumor treatment procedures are usually complicated where tumor detection and preliminary diagnosis are based on magnetic resonance images (MRI), and treatment planning also uses histopathological and molecular analysis of the tissue sample^6^. Diagnosis by radiologists, when comparing the first MRI diagnosis to the final histology diagnosis, varies greatly among tumor types and locations, with an overall accuracy of 72% for broad tumor type classification, which shows the need for computational methods to improve qualitative assessments^7^. With recent technological advances, deep learning algorithms can be trained to assist radiologists in diagnosing brain tumors based on MR images. Even though deep learning methods have led to reasonable advancements in adult brain tumor detection, classification and segmentation^8–10^, their implementation in pediatric cases has been limited^11,12^ mainly due to the lack of large and standardized open access datasets^13,14^. Deep learning models trained on MR images from adults will not perform well on images from children, since PBTs have different diagnostic properties. The “Children’s Brain Tumor Network” (CBTN)^15,16^ is one of the largest PBT datasets, and could potentially be used in the future similarly to the adult brain tumor segmentation challenge (BraTS)^17–19^, as a standard and reference dataset for development and comparison of deep learning methods^20^. This study is, to the best of our knowledge, the first report on the implementation of deep learning on the MR dataset from CBTN for brain tumor classification, and also one of few hitherto published MR image-based deep learning studies on any brain tumor pediatric dataset.

This work aimed to investigate methods for achieving a reliable and explainable multiclass classification of PBTs, considering different MRI sequences, and fusing age and image information. Several convolutional neural networks (CNNs) were implemented and evaluated using model performance metrics, as well as visualization and quantification of model activation maps.

## Material and Methods

### Dataset cohort

The dataset was obtained upon application to and approval from CBTN^15^ (accessed in 2021). Only the six largest tumor categories were acquired from the CBTN source data^15^. These categories contained 273 subjects and 789 examination sessions, where during a session one or more scans with different MR sequences could have been taken. Here, a session refers to each time a patient underwent an MRI examination, *i*.*e*., a T2-w, T1-w, T1w-Gd, FLAIR, and/or diffusion-weighted (DW) scan. Subject ages ranged from two months to 18 years old. The tumor types available in the original dataset were low-grade astrocytoma (ASTR) (132 subjects with 389 sessions), medulloblastoma (MB) (67 subjects with 199 sessions), ependymoma (EP) (45 subjects with 127 sessions), atypical teratoid rhabdoid tumor (ATRT) (20 subjects with 55 sessions), diffuse intrinsic pontine glioma (DIPG) (six subjects with 14 sessions), ganglioma (one subject with one session), germinoma (one subject with two sessions), and teratoma (one subject with two sessions).

### Data selection and exclusion

An automatic selection was implemented on the original dataset based on the quality of the scans, and subsequently on visual assessment of the scans that passed the first filtering. Quality selection was based on the voxel resolution, filtering out images with less than 3 mm in resolution in the longitudinal and coronal directions, and with less than 10 transversal slices. These values were chosen to avoid artefacts due to a low image resolution. In contrast, for DW images the selection was instead based on the availability of at least six diffusion encoding directions used for computing an ADC-map. Visual assessment of all images in each scan excluded a scan if: (i) the tumor was not visible, (ii) image artifacts (motion, metal, induced by neurosurgical clips) were present, (iii) the transversal plane had been clipped, (iv) images were acquired post-operatively, and (v) images showed the spine only. By visual assessment, the tumor location was saved as boundary box coordinates identifying the tumor region, using ITK Snap^21^. The age of the patient at the time of the scan was also used in the analysis.

### Data preprocessing

#### Normalization and re-sampling

Data harmonization was performed because the CBTN dataset was collected on a variety of MRI scanners^16^. The brain from each MRI volume was extracted using a deep learning-based brain extraction tool^22^, followed by per-sequence voxel intensity normalization using the 0.2% percentile minimum and maximum intensity values. Lastly, each volume was isotropically interpolated to 1 mm isotropic resolution using an order five spline interpolation function (`nibabel.processing.conform`). The final volumes were then reshaped to have 240 × 240 pixels in the transverse plane. Tumor boundary boxes were re-scaled to match the final scans’ resolution and size.

#### ADC-computation

The DW-MR data was processed using MRtrix3 software^23^ to obtain a diffusion tensor from which the ADC map was calculated. Briefly, Gibbs-ringing artefact removal, denoising and motion correction were performed before the diffusion tensor fitting. The ADC map was then computed.

#### Tumor slice selection

The extracted transversal 2D slices and the location of the slice within the boundary box of the tumor region were saved. The slice position was used to select the slices positioned within 20-80% of the tumor boundary box. This choice ensures that images showing only small portions of the tumor were not included. Transversal slices were used instead of the volumetric data due to the limited number of subjects available for training a 3D CNN model.

### Age as input

Age information was extracted from the NiFTi file name. The age information was the age in days of the subject on the day when the earliest scan was taken. All ages were converted into age in years and normalized within each tumor class in the range of [0, 1] by subtracting the minimum and dividing the result by the difference between the maximum and the minimum. The final composition of the dataset with age and sex information is summarized in Table 1.

**Table 1.** Subject-wise summary of the dataset with tumor location and scans available for each MR sequence. The age is averaged for all the sequences used. For subjects with multiple sessions, the age information was obtained from the earliest scan available. m: mean, sd: standard deviation, M: male, F: female, NA: not available.

| Tumor type | Subjects (infra/supra/both) | Sex (M/F/NA) | Age in years<br>median<br>[range]<br>(m ± std) | Subjects Scans<br>Extracted slices<br>(infra/supra/both) | | |
| --- | --- | --- | --- | --- | --- | --- |
|  |  |  |  | T2-w | T1w-Gd | ADC |
| ASTR | 92<br>(39/50/3) | 53/39/0 | 7.61<br>[0.53, 17.40]<br>7.78 ± 4.57 | 33/41/2<br>44/63/6<br>729/967 | 31/43/3<br>41/57/9<br>615/792 | 16/19/3<br>16/22/3<br>287/439 |
| EP | 30<br>(18/9/3) | 19/11/0 | 3.74<br>[0.00, 17.98]<br>6.02 ± 5.00 | 17/9/3<br>19/14/10<br>394/434 | 17/7/3<br>20/11/9<br>367/377 | 5/3/1<br>6/3/2<br>178/109 |
| MB | 59<br>(59/0/0) | 33/22/4 | 9.04<br>[0.24, 17.86]<br>8.77 ± 4.51 | 52/0/0<br>57/0/0<br>1000/0 | 49/0/0<br>57/0/0<br>790/0 | 19/0/0<br>19/0/0<br>370/0 |
| **Total** | 181<br>(116/59/6) | 105/72/4 | 7.56<br>[0.00, 17.98]<br>7.81 ± 4.71 | 102/50/5<br>120/77/16<br>2123/1401 | 97/50/6<br>118/68/18<br>1808/1169 | 40/22/4<br>41/25/5<br>835/548 |

### Data splitting

Scans were split subject-wise, into training and validation (80%) and testing (20%), ensuring that subjects with multiple scans did not end up in the same set to avoid data leakage^24,25^. Five times repeated ten-fold stratified cross-validation was employed in all experiments to account for the small size of the dataset and to make sure that all data could be used for training, validation, and testing in the different repetitions.

### Data augmentation

To account for the variation of image types in the dataset (including orientation and brightness), data augmentation was applied^26–29^, namely random rotations between ±45°, random increase in brightness up to 50%, random horizontal and vertical flipping, and 10% random width and height shift.

#### Slice and volume class prediction

To obtain a per-volume classification, predictions were first computed for each slice in a volume and then aggregated into a single prediction for the whole volume. The classification of each slice was obtained by leveraging the models trained through the ten-fold stratified cross-validation scheme, ensembling the models’ predictions using two methods: soft voting and hard voting. For soft voting, the class with the highest mean predicted probability over the cross-validation models was assigned as the ensemble prediction for a slice in a volume:

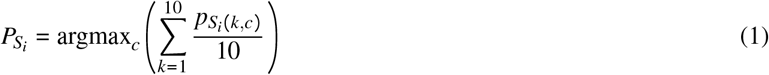

where *S*_*i*_:*i*-th slice in a volume, 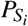: ensemble prediction for *S*_*i*_, *c*: classes, *k*: *k*-th model from the ten-fold stratified cross validation scheme, and 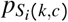: predicted probability for class *c* for *S*_*i*_ from the *k*-th model. For the hard voting, the most recurring class over the k-folds was considered as the ensemble prediction for a slice in the volume:

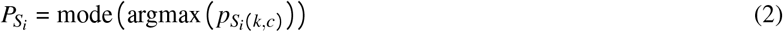

Two methods were then used to aggregate the classification of each slice in a volume-wise tumor type classification: weighted and non-weighted aggregation. In the non-weighted aggregation, the most recurring predicted class over the slices belonging to a volume was taken as a prediction for the whole volume:

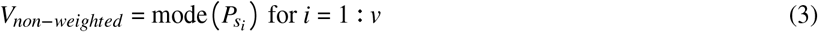

where *v* is the number of slices in the volume, *V*. For the weighted aggregation, a weight, *w*_*i*_, equal to the slice position within the tumor boundary box was assigned to each slice. Slices at the extremities were assigned a lower weight than those near the center of the tumor-bounding box. The class with the highest sum of the weights was saved as the prediction for the volume:

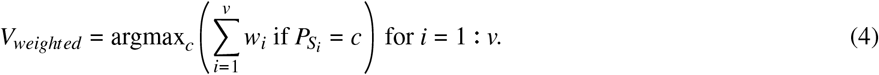

### Network architecture and training

Several deep learning models were investigated including off-the-shelf CNNs, namely visual geometry groups (VGGs) and residual networks (ResNets), and a custom CNN model (2D-SDM4). Models were trained on image data only, as well as on image data combined with age information. In the latter case, CNN models were used to encode the image data, while a tabular network described below encoded the age information. Figure 1 shows a schematic representation of the different models when trained on image and age information. Initial experiments on the final image dataset were carried out to investigate which network to use in the subsequent experiments. The specifications of the different models are given below.

**Figure 1.**
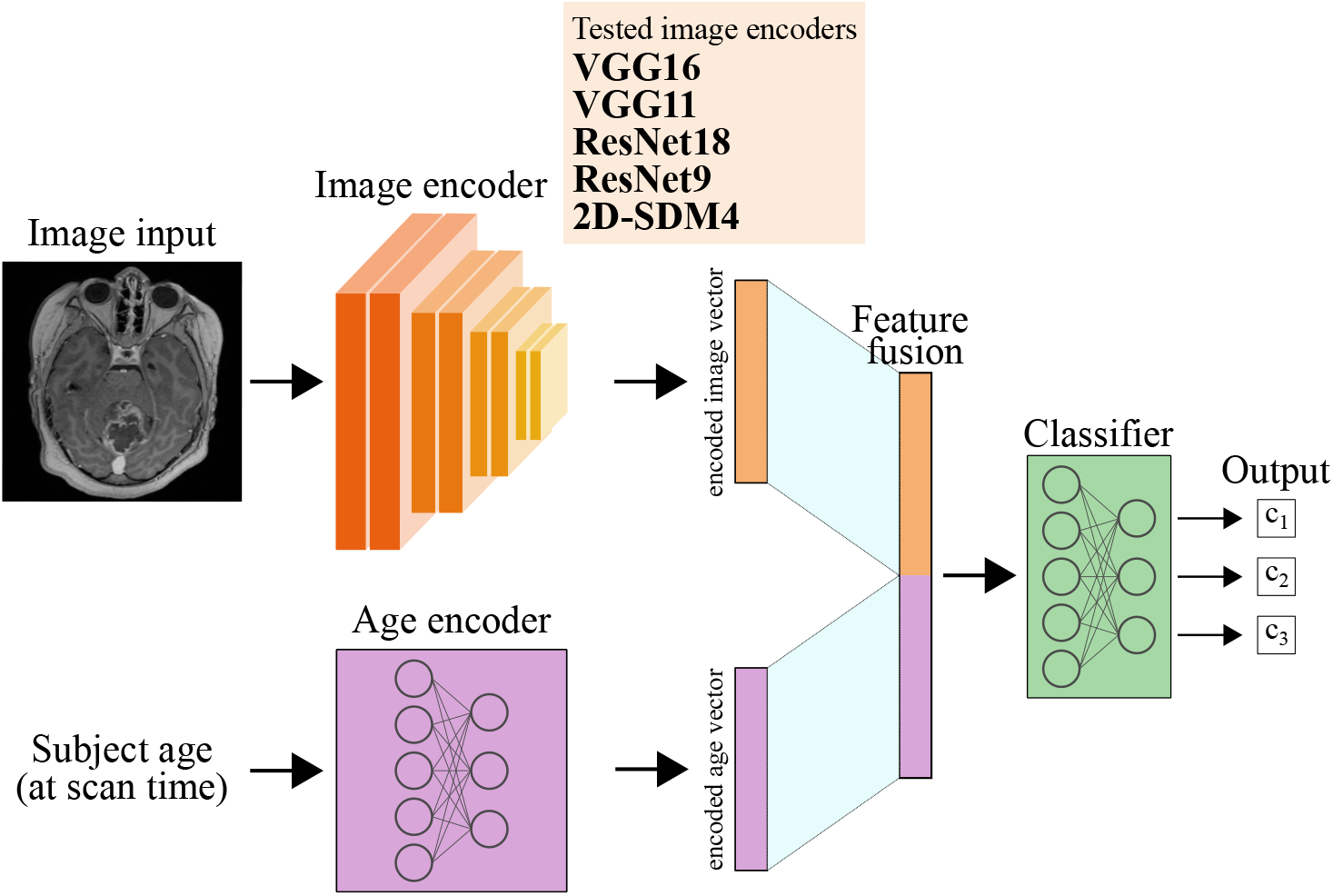
Schematic representation for the image encoders used along with the age encoder and their concatenation that leads to the final classification of each 2D image.

#### Off-the-shelf-models

Among the off-the-shelf 2D CNN models, VGG11 and VGG16^30^, and ResNet9 and ResNet18^31^ were chosen given their extensive use in literature as CNN-based image classifiers, and to allow comparison with previous results.

#### 2D-SDM4 custom model

A simple and shallow detection model with four convolutional blocks (2D-SDM4) is a custom-made network^32^ consisting of 16 layers, of which eight were 2D convolutional layers with a 3x3 kernel and an increasing number of filters from 64 to 512. Compared to VGG16 which has ≈15 million parameters distributed over 13 convolutional layers, 2D-SDM4 has ≈5 million parameters distributed over eight convolutional layers. A detailed description of the 2D-SDM4 model architecture is available in^33^.

#### Image and age data fusion

Several data fusion approaches have been proposed^34–36^, such as early fusion, joint fusion and late fusion. In this work, joint fusion (feature fusion) was used to combine image and age information for the prediction of tumor type. The advantage of joint fusion is that a single model is trained on both image and age information, with the age not blended immediately with the risk of not being properly used^36^. The age was encoded using a tabular network composed of three dense layers with increasing nodes from 8 to 32, and each dense layer was followed by a Rectified Linear Unit (ReLU)^37^ activation and batch normalization layer. The encoded age vector was then concatenated to the encoded image vector before being fed to the classifier part of the model.

#### Model pre-training

All CNN models used in this study were pre-trained from randomly initialized weights on the BraTS2020 dataset^17–19^, instead of natural images, for the task of classifying 2D transversal slices with or without a tumor. Model pre-training was performed on the adult brain tumor dataset because of the limited availability of pediatric data, given the similarity between such datasets. Models were pre-trained on T2-w or T1w-Gd images separately, to match the MR-sequences available for the CBTN dataset. For ADC images the model weights from models trained on T2-w images were used. During the experiments with the pediatric dataset, transfer learning was employed where half of the layers were unfrozen for the pre-trained off-the-shelf networks, whereas for 2D-SDM4 all layers were unfrozen. The weights for the tabular network were randomly initialized and all the layers were trainable.

#### Model training

Training was performed on images of size 240 × 240 pixels and a batch size of 32. Class weights were used during the experiments to account for the imbalance in the dataset, with weights computed on the training dataset. The Adam optimizer^38^ was employed along with LookAhead^39^. The initial learning rate was set to 1e-4 decreasing 1% of its value after every epoch. Models were trained for 100 epochs, without early stopping. When image and age information was used, the image and age encoders were jointly trained.

### Classification tasks

Two classification tasks were defined as: (1) three classes of infratentorial PBT types (medulloblastoma, astrocytoma and ependymoma) and (2) five classes of PBT types where the location was accounted for (medulloblastoma infratentorial, astrocytoma infratentorial, ependymoma infratentorial, astrocytoma supratentorial, ependymoma supratentorial). These tasks were investigated on MR images acquired using T1w-Gd, T2-w and ADC sequences.

### Evaluation metrics and statistical methods

To evaluate the performances of models on the unbalanced dataset, Matthew’s correlation coefficient (MCC) [-1, 1] was computed^40–42^. MCC equal to +1 indicates a perfect classification, whereas MCC equal to 0 is equivalent to random chance classification^40^. Accuracy was computed to allow comparison of the results with other works, which is a more commonly used metric but is not suitable for an unbalanced dataset. To get a view of how the model performed in each class, F1 score, precision, and recall were computed class-wise. The overall F1 score was calculated using macro-averaging. Random chance for the different classification tasks was computed using 10,000 random permutations over a dummy dataset containing the same number of samples for each class. The Wilcoxon signed-rank test was used to investigate if the fusion of age information significantly improved model performance, by comparing models trained on T2-w images alone or fused with age. A p-value < 0.05 was considered significant.

### Model explainability

In this work, model explainability was used to provide a visual explanation and a validation of the image regions used for classification. Given that the tumor is the most important feature when performing the diagnosis, the model should be able to focus as much as possible on the tumor. Grad-CAM^43,44^ is a type of post-hoc attribution method used to visualize the most important features in an image (activation maps) when predicting a class for a selected layer. In this work, the Grad-CAM activation maps were thresholded at the 95% percentile of their intensity to highlight the most important features in the image used for classification. Additionally, the percentage of the overlap between the thresholded activation maps and the tumor region boundary box was computed. To test if the model was able to focus on the tumor consistently, augmentation was performed as ten different rigid transformations (flipping and rotation) that were randomly applied to each test image. Due to rescaling, some error was expected in the position of the boundary boxes which could affect the calculated overlap area. Activation maps were computed for each transformed image along with the percentage of their overlap with the tumor region.

## Results

From a preliminary investigation, among the four off-the-shelf networks, the VGG16 model performed best on task 1 (image: MCC = 0.52 ± 0.17, accuracy = 0.71 ± 0.10, image+age: MCC = 0.55 ± 0.15, accuracy = 0.72 ± 0.09). The custom-made network when trained on image data only obtained the highest performance, while having a smaller number of model parameters. The age information alone was not predictive of the tumor classes. Moreover, no improvements in model performance could be seen when the image was fused with age using a shallow age encoder (1 dense layer) or no encoder. Therefore, results are presented in detail for the 2D-SDM4 model for all the classification tasks as well as for model explainability.

### Classification of the infratentorial tumors

The results of the 2D-SDM4 model performance on task 1 with and without age information, using three different MR sequences of T1w-Gd, T2-w and ADC-images are summarized in Table 2. The weighted soft voting slice aggregation method gave the best results for volume-wise classification. The metrics (MCC and F1 score) were highest for the ADC sequence. In all cases, the performance for the EP class was lower compared to the other two tumor types. Combination of T2-w with the patient’s age improved the performance metrics, (p-value < 0.05 for MCC values). Combination of ADC sequences with age however did not improve the performance metrics (p-value > 0.05 for MCC values). Sankey diagrams for the performance on task 1 when using image and age information combined are shown in Figure 2.A for T2-w images and in Figure 2.C for ADC.

**Table 2.**
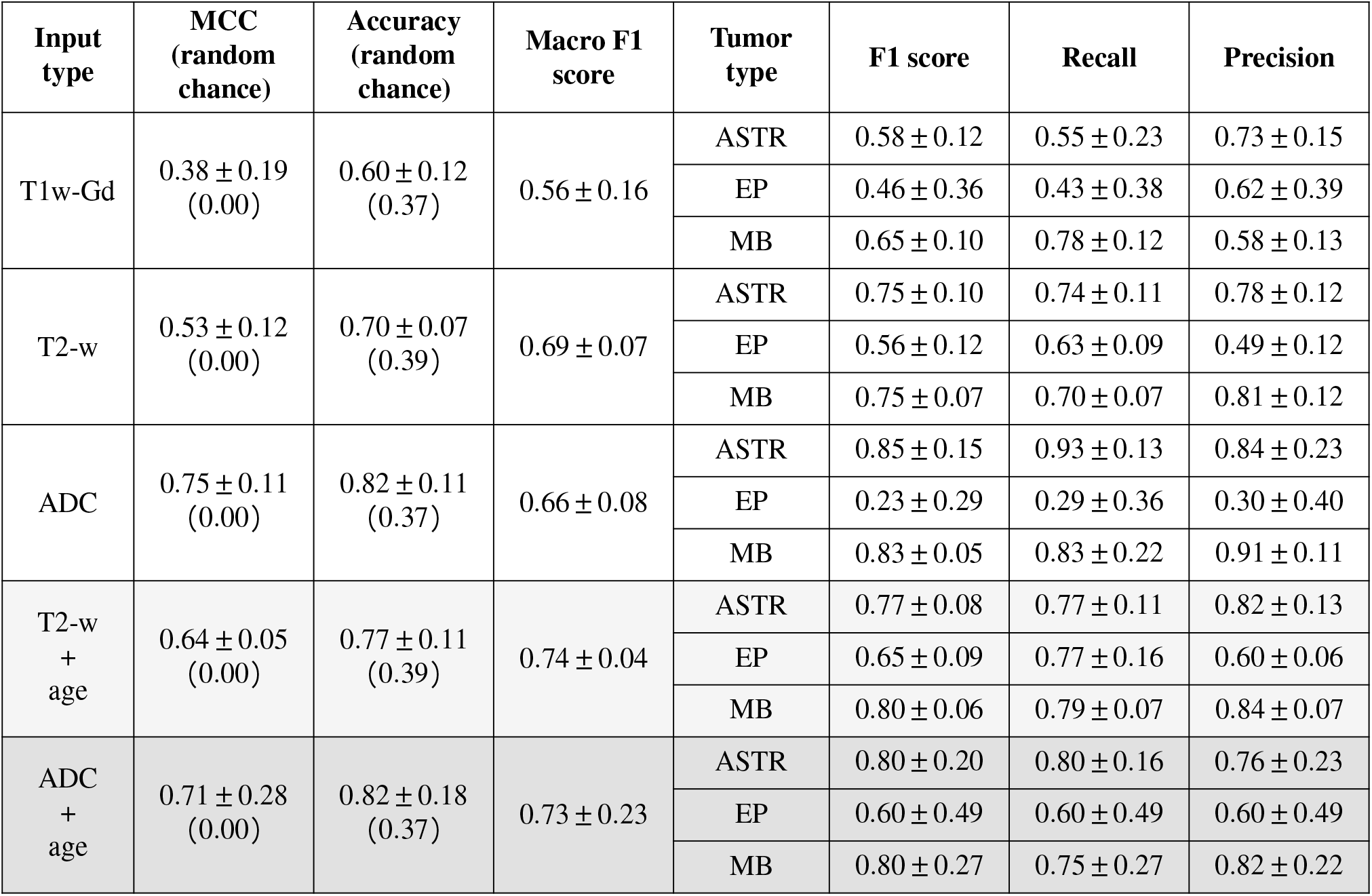
Results of 2D-SDM4 network performance for the comparison of MR sequence alone and fused with the patient age for the two best-performing MR sequences. Weighted soft voting provided the highest volume-wise classification.

**Figure 2.**
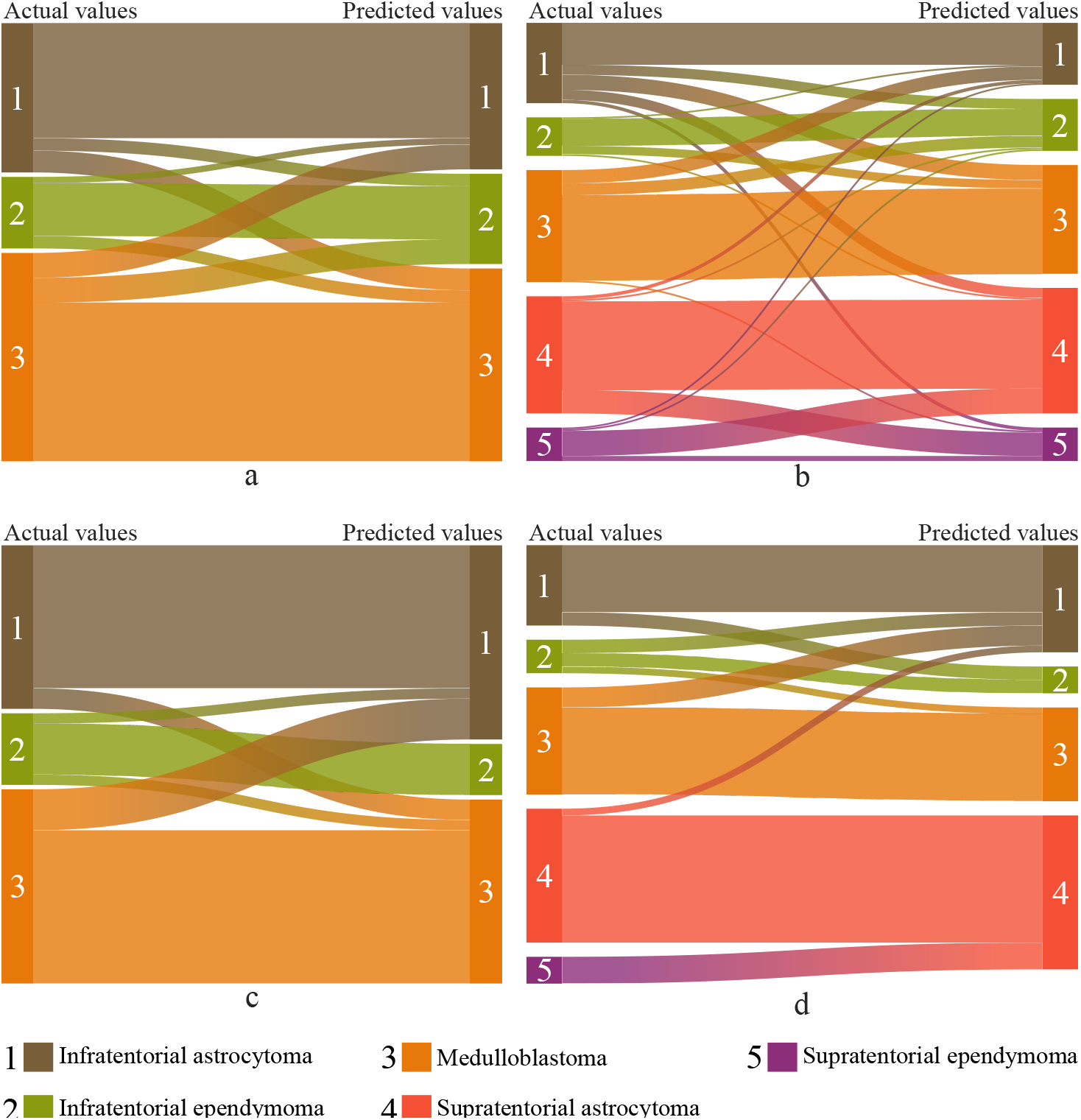
Sankey diagrams showing the flows for the predictions of T2-w (top row) and ADC (bottom row) images for task 1 with age, solely infratentorial images (a, c) and task 2 with age, supratentorial and infratentorial (b, d). The flows are proportional to the average number of scans in the test sets used in the repeated cross-validation scheme.

### Classifcation of infra- and supratentorial tumors

The results for task 2 with and without the age information for the classification using blended infratentorial and supratentorial tumor volumes, are shown in Table 3. The results from the non-weighted hard voting were the best for volume-wise classification of T2-w scans and non-weighted soft voting were the best for volume-wise classification of ADC scans. The fusion of image and patient’s age significantly improved the model MCC performance from 0.49 to 0.57 when using T2-w (p-value < 0.05). Sankey diagrams presenting the performance of task 2 for T2-w images are shown in Figure 2.B and for ADC are shown in Figure 2.D.

**Table 3.** Results from the classification of supratentorial and infratentorial PBTs using image and age information combined. Non-weighted hard voting provided the highest metrics for T2-w images, and non-weighted soft voting provided the highest metrics for ADC images. Results are presented with their mean and standard deviations over a five-repetition ten-fold stratified cross-validation. m: mean, sd: standard deviation.

| Input type | MCC (random chance) | Accuracy (random chance) | Macro F1 score | Tumor type | F1 score | Recall | Precision |
| --- | --- | --- | --- | --- | --- | --- | --- |
| T2-w + age | $0.57 \pm 0.05$<br>(0.00) | $0.66 \pm 0.04$<br>(0.24) | $0.57 \pm 0.05$ | ASTR infra | $0.58 \pm 0.22$ | $0.52 \pm 0.29$ | $0.68 \pm 0.13$ |
| | | | | ASTR supra | $0.74 \pm 0.10$ | $0.83 \pm 0.08$ | $0.71 \pm 0.18$ |
| | | | | EP infra | $0.58 \pm 0.07$ | $0.60 \pm 0.10$ | $0.52 \pm 0.06$ |
| | | | | EP supra | $0.18 \pm 0.17$ | $0.16 \pm 0.15$ | $0.23 \pm 0.20$ |
| | | | | MB | $0.78 \pm 0.06$ | $0.73 \pm 0.09$ | $0.78 \pm 0.04$ |
| ADC + age | $0.70 \pm 0.09$<br>(0.00) | $0.77 \pm 0.07$<br>(0.25) | $0.57 \pm 0.1$ | ASTR infra | $0.72 \pm 0.09$ | $0.83 \pm 0.17$ | $0.68 \pm 0.19$ |
| | | | | ASTR supra | $0.89 \pm 0.02$ | $0.96 \pm 0.06$ | $0.82 \pm 0.02$ |
| | | | | EP infra | $0.42 \pm 0.43$ | $0.50 \pm 0.50$ | $0.38 \pm 0.41$ |
| | | | | EP supra | $0.00 \pm 0.00$ | $0.00 \pm 0.00$ | $0.00 \pm 0.00$ |
| | | | | MB | $0.85 \pm 0.15$ | $0.81 \pm 0.21$ | $0.94 \pm 0.11$ |

### Grad-CAMs

Representative Grad-CAMs computed on test images using the 2D -SDM4 model trained on T2-w and ADC data are shown in Figure 3 and Figure 4, for non-augmented and augmented test images, respectively. Moreover, Table 4 shows the quantification of the Grad-CAMs overlapping with the tumor region. Activation maps for 2D-SDM4 covered a larger tumor region compared to VGG16, whose results are not shown here. A similar trend could be observed when randomly augmenting the test images. Results when considering the non-augmented test images show a higher coverage than when randomly augmenting the images. One reason could be that by changing the location of the tumor in the image, it becomes more difficult for the network to identify the tumor, indicating that data augmentation did not work as much as expected.

**Table 4.** Results for the computation of the tumor region covered by the class activation map with augmentation and without augmentation of the test images. m: mean, sd: standard deviation.

| Network | Augmentation of test images | Heatmap area over tumor ROI (m±sd) | Heatmap area over tumor ROI (median, range) |
| --- | --- | --- | --- |
| 2D-SDM4 on T2-w images | No | $0.49 \pm 0.26$ | 0.48<br>[0.00, 1.00] |
| | Yes | $0.28 \pm 0.17$ | 0.26<br>[0.00, 0.95] |
| 2D-SDM4 on ADC images | No | $0.42 \pm 0.23$ | 0.43<br>[0.00, 0.93] |
| | Yes | $0.24 \pm 0.17$ | 0.22<br>[0.00, 0.64] |

**Figure 3.**
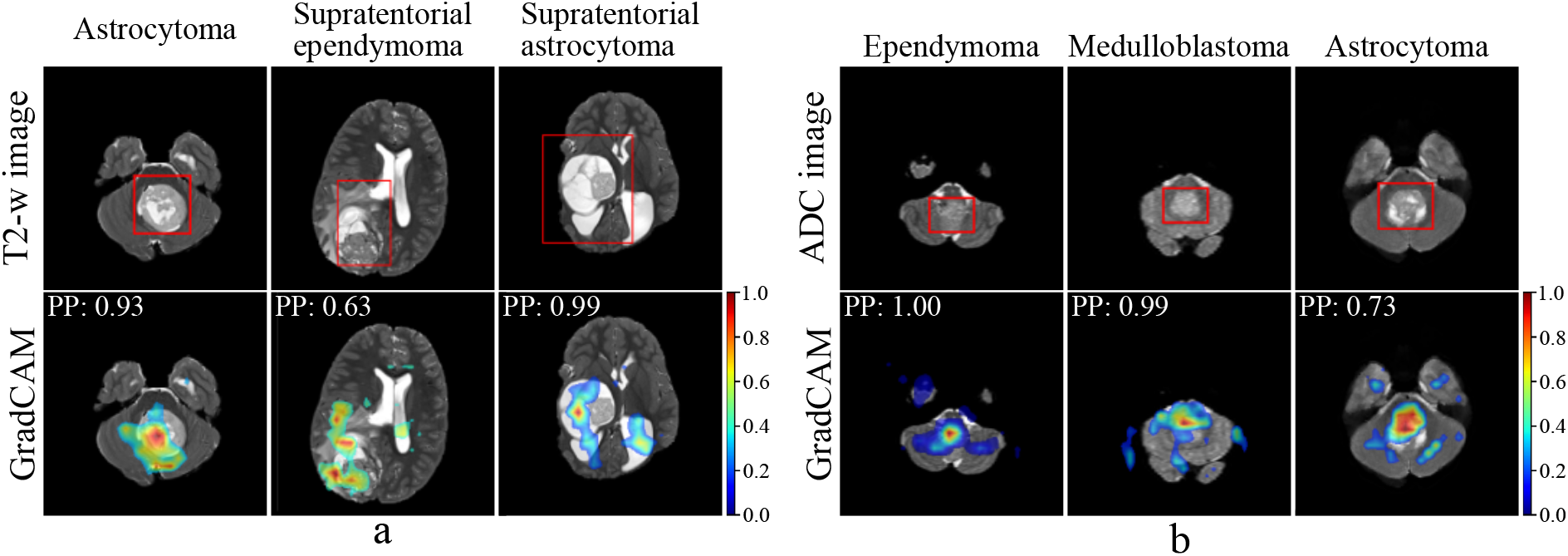
Grad-CAMs for different classes on T2-w images (a) and ADC images (b) obtained from network 2D-SDM4 on tasks 1 and 2 with age information. The model’s prediction is equal to the ground truth for all the images presented. The predicted probability (PP) is also reported.

**Figure 4.**
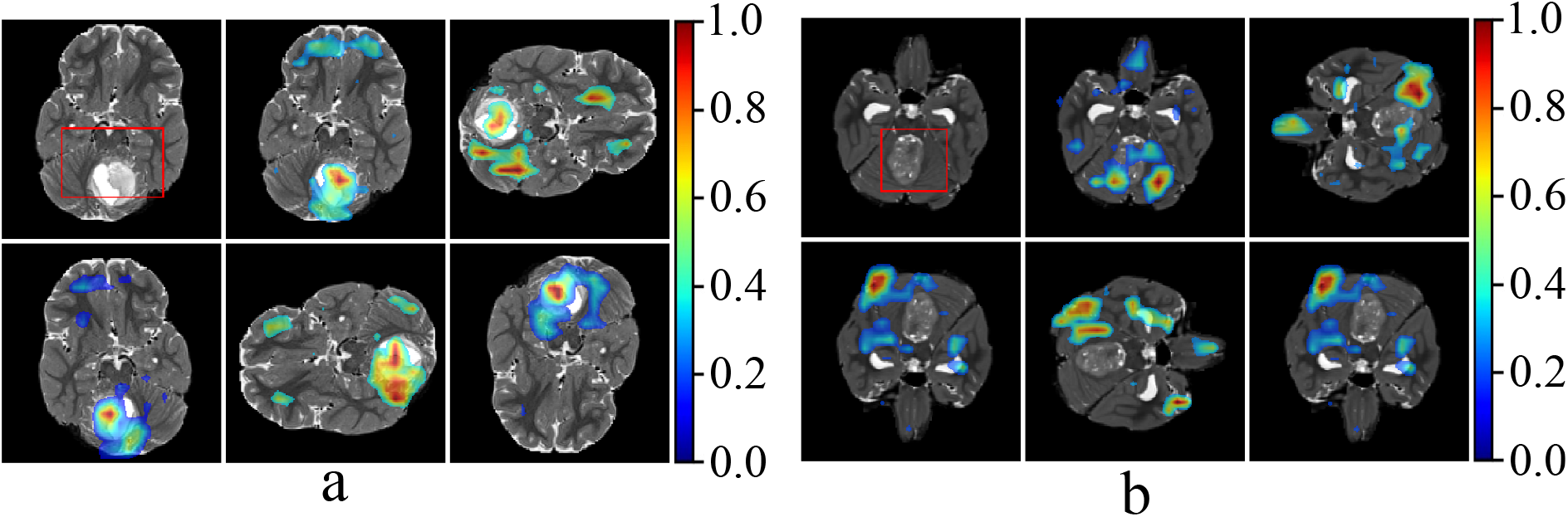
Examples of Grad-CAM visualization for 2D-SDM4 network of randomly augmented T2-w test images of a medulloblastoma in which the model was able (a) and was not able (b) to follow the tumor as the image was rotated. For both cases, the model prediction was equal to the ground truth.

## Discussion

In this study, deep learning methods were implemented and evaluated for the classification of PBT types based on MR images from the CBTN dataset. The effect of network architectures, input image types, addition of patient age and location of the tumor on classification performance were investigated. The evaluation was based on the performance metrics, and the visual and quantified model explanatory maps.

### Network architecture

ResNets and VGGs were implemented in this study inspired by previous reports in the literature for similar tasks on both pediatric^11,12^ and small adult brain tumor datasets^45,46^. However, instead of using ResNet50^11,12^, ResNet18 and ResNet9 were implemented since shallower networks have been reported as being more suitable for small datasets^47^. The custom shallow network 2D-SDM4 was the best-performing network, which agreed with previous reports^26,48–50^. The fewer parameters used in shallow networks appear to generalize the information obtained from small datasets more efficiently compared to more complex networks, for which small datasets do not provide sufficient information. Moreover, when looking at model explainability, Grad-CAM-derived activation maps for the shallower network showed a higher degree of overlap with the tumor boundary box compared to the VGG16 model, for both augmented and non-augmented test images.

### MR image sequences

To test the concept of multimodal MR inputs, T2-w and T1w-Gd images were registered and used as input for a simple classification task using early fusion which did not result in any improvement in the performance of the networks compared to using a single MR sequence image. Other combinations of MR sequences were not tested since not all subjects had been examined using all types of MR-sequences. Moreover, Artzi *et al*.,^12^ and Quon *et al*.,^11^ both tested combinations of three different MR sequences and in both cases these combinations unexpectedly led to underperformance, compared to single MR sequence inputs. Artzi *et al*., registered the MR-sequences to obtain a single input, but Quon *et al*., did not mention details of the implemented combination methods. Given that only early fusion was used in all of these works to combine the different MRI sequences, the joint and/or late fusion approaches should also be investigated.

Among the single MR-sequences, T2-w images achieved more consistent class-wise metrics, especially for ependymomas, whereas the overall highest accuracy and MCC were obtained using ADC images. The performance of models trained on T2-w images showed less variation over the repeated cross-validations, and was in accordance with Quon’s results^11^. Moreover, even though MCC and accuracy values for models trained on T2-w data were slightly lower compared to when ADC data was used, class-wise metrics for EP were higher in the case of T2-w explained by the larger number of images.

The results on ADC data were in accordance with those of Artzi *et al*.,^12^. However, in our study, only 1383 ADC images were available with a high class imbalance and with EPs presenting only seven scans to split between training and testing. This resulted in a large variation in model performance over the repeated cross-validations, even though the class-wise metrics for ASTR and MB were the highest compared to the other MR sequences tested.

### Location of the tumors

The study investigated the classification of infratentorial tumors and also supratentorial tumors. The latter have not been reported earlier. The results for the supratentorial tumors obtained in this study show the potential of deep learning models trained on MRI data for supporting radiologists and improving the diagnosis of pediatric supratentorial tumors which have been shown to be hard to classify^7^. It should however be considered that the supratentorial categorization of the tumors is slightly different in these two studies. Our results also agree with the report by Dixon *et al*.,^7^ in that tumors in the posterior fossa were easier to diagnose.

### Age information

The fusion of T2-w image and age data showed an improvement in MCC (p-value < 0.05) and accuracy compared to when only image data was used for both of the 3- and 5-classes tasks. This suggests that additional information relevant to the specific class can help networks improve performance when images might not be providing sufficient class-specific features. In this work, a joint fusion approach was used to combine the image and age information during model training, allowing the image and age encoder to be jointly trained. Moreover, the results also showed that the depth of the age encoder has to be considered. In fact, only when the age information was encoded using the three dense layers did the joint fusion approach improve classification performance.

### Model explainability

CNNs have been used for a wide range of tasks in medical image analysis. Clinical usage requires a reliable^51^ explainability of the decision process behind a network’s predictive power. To assess the amount of information used from the tumor region, the area covered by the class activation maps in relation to the tumor region defined by a boundary box was computed showing an overall 48% activation overlapping the tumor region. Overall, in shallower networks spatial information is better preserved compared to deeper models that learn more abstract features while losing spatial resolution due to the pooling layers. Therefore, model explainability based on the localization of areas with high activation and consequent measurement of overlap between Grad-CAM and the tumor region of interest, allows shallow networks to obtain better explainability.

### Comparison with related work

Quon *et al*.,^11^ reported an F1 score of 0.80 (accuracy of 0.92) and Artzi *et al*., 0.82 (accuracy of 0.87) compared to a macro F1 score of 0.74 (accuracy of 0.81) for ADC and 0.73 (accuracy of 0.77) for T2-w in this study, considering the infratentorial tumors. None of the previous studies specified the averaging method for the F1 score. However, Artzi *et al*.,^12^ reported the class-wise F1 scores (ASTR: 0.87 ± 0.01, EP: 0.41 ± 0.06, MB: 0.82 ± 0.03) for their best-performing sequence, *i*.*e*. Trace/ADC with age and Quon *et al*., reported approximate values (ASTR: 0.8, EP: 0.40, MB: 0.8) for T2-w images. The class-wise results for ASTR and MB are comparable to our study. EP had lower scores in all studies; however, the score was higher in our study (by ≈ 0.2), which might be due to compensation for the imbalances present in EPs through the use of class weights during the experiments. It should be noted that the comparison of the performance metrics among the studies should be considered in general terms due to the differences in the datasets and the number of classes for the defined tasks. The reliability of the results is also affected by the experimental protocols, as Quon *et al*.,^11^ applied a five-fold stratified cross-validation and employed the top five models and Artzi *et al*.,^12^ implemented a five-fold stratified cross-validation to compute the final predictions. The work presented here, unlike the two papers mentioned, employed the entire dataset and tested the network on all subjects in the dataset by implementing five repetitions of a ten-fold stratified cross-validation to achieve the most consistent and reliable results possible.

### Limitations

There are some limitations in this work, primarily regarding the dataset but also the methods employed. The dataset, although the largest accessible, has a relatively small size and the presence of the different tumor types was quite unbalanced after filtering. Not all tumor types originally available could be taken into consideration, which reduced the number of classes. Moreover, the quality of the scans overall varied greatly between each scan. Further investigations could still be carried out, especially regarding the tabular network whose architecture can be further improved.

## Conclusions

In this study, the classification of PBTs based on MR images from the CBTN dataset was reasonably well achieved using deep learning methods, which could aid radiologists in the assessment of these cases. The shallow network trained on fused image and age information had higher performance for the classification tasks implemented on the relatively small dataset in this study. Among the MR sequences, T2-w and ADC were the most suitable ones for tumor type classification. Future studies with larger datasets, the fusion of multiple MR-sequences and extended clinical information along with further refinements of the network designs are warranted.

## Data Availability

All data produced are available online at https://cbtn.org/

## Acknowledgments

The research was made possible in part due to The Children’s Brain Tumor Tissue Consortium (CBTTC) / The children’s brain tumor network (CBTN). The study was financed by Swedish Childhood Cancer Foundation (MT2021-0011, MT2022-0013), Joanna Cocozza’s Foundation (2023-2024), Linköping University’s Cancer Strength Area (2022) and ALF Grants, Region Östergötland (974566).

## References

1. Ferlay, J. et al. Global cancer observatory: cancer today. https://gco.iarc.fr/today/home (2020). Accessed: 2022.

2. Sharma, R. A systematic examination of burden of childhood cancers in 183 countries: estimates from GLOBOCAN 2018. Eur. J. Cancer Care 30, e13438 (2021).

3. Ostrom, Q. T. et al. CBTRUS statistical report: primary brain and other central nervous system tumors diagnosed in the United States in 2013–2017. Neuro-oncology 22, iv1–iv96 (2020).

4. Corti, C. et al. Effects of supratentorial and infratentorial tumor location on cognitive functioning of children with brain tumor. Child’s Nerv. Syst. 36, 513–524 (2020).

5. Pollack, I. F. Brain tumors in children. New Engl. J. Medicine 331, 1500–1507 (1994).

6. Louis, D. N. et al. The 2021 WHO classification of tumors of the central nervous system: a summary. Neuro-oncology 23, 1231–1251 (2021).

7. Dixon, L., Jandu, G. K., Sidpra, J. & Mankad, K. Diagnostic accuracy of qualitative MRI in 550 paediatric brain tumours: evaluating current practice in the computational era. Quant. Imaging Medicine Surg. 12, 131 (2022).

8. Ali, S. et al. A comprehensive survey on brain tumor diagnosis using deep learning and emerging hybrid techniques with multi-modal MR image. Arch. Comput. Methods Eng. 29, 4871–4896 (2022).

9. Amin, J., Sharif, M., Haldorai, A., Yasmin, M. & Nayak, R. S. Brain tumor detection and classification using machine learning: a comprehensive survey. Complex & Intell. Syst. 1–23 (2021).

10. Tandel, G. S. et al. A review on a deep learning perspective in brain cancer classification. Cancers 11, 111 (2019).

11. Quon, J. et al. Deep learning for pediatric posterior fossa tumor detection and classification: a multi-institutional study. Am. J. Neuroradiol. 41, 1718–1725 (2020).

12. Artzi, M. et al. Classification of pediatric posterior fossa tumors using convolutional neural network and tabular data. IEEE Access 9, 91966–91973 (2021).

13. Shaari, H. et al. Deep learning-based studies on pediatric brain tumors imaging: narrative review of techniques and challenges. Brain Sci. 11, 716 (2021).

14. Huang, J., Shlobin, N. A., Lam, S. K. & DeCuypere, M. Artificial intelligence applications in pediatric brain tumor imaging: A systematic review. World neurosurgery 157, 99–105 (2022).

15. The Children’s Brain Tumor Network. https://cbtn.org/. Accessed: 2021.

16. Lilly, J. V. et al. The children’s brain tumor network (CBTN)-Accelerating research in pediatric central nervous system tumors through collaboration and open science. Neoplasia 35, 100846 (2023).

17. Menze, B. H. et al. The multimodal brain tumor image segmentation benchmark (BRATS). IEEE transactions on medical imaging 34, 1993–2024 (2014).

18. Bakas, S. et al. Advancing The Cancer Genome Atlas glioma MRI collections with expert segmentation labels and radiomic features. scientific data 4 (1), 170117 (2017).

19. Bakas, S. et al. Identifying the best machine learning algorithms for brain tumor segmentation, progression assessment, and overall survival prediction in the BRATS challenge. preprint arXiv:1811.02629 (2018).

20. Fathi Kazerooni, A. et al. Automated Tumor Segmentation and Brain Tissue Extraction from Multiparametric MRI of Pediatric Brain Tumors: A Multi-Institutional Study. Neuro-Oncology Adv. vdad027 (2023).

21. Yushkevich, P. A., Gao, Y. & Gerig, G. ITK-SNAP: An interactive tool for semi-automatic segmentation of multi-modality biomedical images. In 2016 38th annual international conference of the IEEE engineering in medicine and biology society (EMBC), 3342–3345 (IEEE, 2016).

22. Isensee, F. et al. Automated brain extraction of multisequence MRI using artificial neural networks. Hum. brain mapping 40, 4952–4964 (2019).

23. Tournier, J.-D. et al. MRtrix3: A fast, flexible and open software framework for medical image processing and visualisation. Neuroimage 202, 116137 (2019).

24. Yagis, E. et al. Effect of data leakage in brain MRI classification using 2D convolutional neural networks. Sci. reports 11, 22544 (2021).

25. Tampu, I. E., Eklund, A. & Haj-Hosseini, N. Inflation of test accuracy due to data leakage in deep learning-based classification of OCT images. Sci. Data 9, 580 (2022).

26. Paul, J. S., Plassard, A. J., Landman, B. A. & Fabbri, D. Deep learning for brain tumor classification. In Medical Imaging 2017: Biomedical Applications in Molecular, Structural, and Functional Imaging, vol. 10137, 253–268 (SPIE, 2017).

27. Chlap, P. et al. A review of medical image data augmentation techniques for deep learning applications. J. Med. Imaging Radiat. Oncol. 65, 545–563 (2021).

28. Kumar, R. L., Kakarla, J., Isunuri, B. V. & Singh, M. Multi-class brain tumor classification using residual network and global average pooling. Multimed. Tools Appl. 80, 13429–13438 (2021).

29. Sajjad, M. et al. Multi-grade brain tumor classification using deep CNN with extensive data augmentation. J. computational science 30, 174–182 (2019).

30. Simonyan, K. & Zisserman, A. Very deep convolutional networks for large-scale image recognition. preprint arXiv:1409.1556 (2014).

31. He, K., Zhang, X., Ren, S. & Sun, J. Deep residual learning for image recognition. In Proceedings of the IEEE conference on computer vision and pattern recognition, 770–778 (2016).

32. Tampu, I. E., Haj-Hosseini, N., Blystad, I. & Eklund, A. Deep learning for quantitative mri brain tumor analysis. medRxiv 10.1101/2023.03.21.23287514 (2023).

33. Tampu, I. E., Haj-Hosseini, N., Blystad, I. & Eklund, A. Deep learning for quantitative mri brain tumor analysis. medRxiv 2023–03 (2023).

34. Acosta, J. N., Falcone, G. J., Rajpurkar, P. & Topol, E. J. Multimodal biomedical AI. Nat. Medicine 28, 1773–1784 (2022).

35. Kline, A. et al. Multimodal machine learning in precision health: A scoping review. npj Digit. Medicine 5, 171 (2022).

36. Huang, S.-C., Pareek, A., Seyyedi, S., Banerjee, I. & Lungren, M. P. Fusion of medical imaging and electronic health records using deep learning: a systematic review and implementation guidelines. NPJ digital medicine 3, 136 (2020).

37. Agarap, A. F. Deep learning using rectified linear units (ReLU). preprint arXiv:1803.08375 (2018).

38. Kingma, D. P. & Ba, J. Adam: A method for stochastic optimization. preprint arXiv:1412.6980 (2014).

39. Zhang, M., Lucas, J., Ba, J. & Hinton, G. E. Lookahead optimizer: k steps forward, 1 step back. Adv. neural information processing systems 32 (2019).

40. Chicco, D. & Jurman, G. The advantages of the Matthews correlation coefficient (MCC) over F1 score and accuracy in binary classification evaluation. BMC genomics 21, 1–13 (2020).

41. Powers, D. M. Evaluation: from precision, recall and F-measure to ROC, informedness, markedness and correlation. preprint arXiv:2010.16061 (2020).

42. Baldi, P., Brunak, S., Chauvin, Y., Andersen, C. A. & Nielsen, H. Assessing the accuracy of prediction algorithms for classification: an overview. Bioinformatics 16, 412–424 (2000).

43. Zhou, B., Khosla, A., Lapedriza, A., Oliva, A. & Torralba, A. Learning deep features for discriminative localization. In Proceedings of the IEEE conference on computer vision and pattern recognition, 2921–2929 (2016).

44. Selvaraju, R. R. et al. Grad-CAM: Visual explanations from deep networks via gradient-based localization. In Proceedings of the IEEE international conference on computer vision, 618–626 (2017).

45. Chelghoum, R., Ikhlef, A., Hameurlaine, A. & Jacquir, S. Transfer learning using convolutional neural network architectures for brain tumor classification from MRI images. In Artificial Intelligence Applications and Innovations: 16th IFIP WG 12.5 International Conference, AIAI 2020, Neos Marmaras, Greece, June 5–7, 2020, Proceedings, Part I 16, 189–200 (Springer, 2020).

46. Rehman, A., Naz, S., Razzak, M. I., Akram, F. & Imran, M. A deep learning-based framework for automatic brain tumors classification using transfer learning. Circuits, Syst. Signal Process. 39, 757–775 (2020).

47. Bianchessi, T. Pediatric Brain Tumor Type Classification in MR Images Using Deep Learning. http://urn.kb.se/resolve?urn=urn:nbn:se:liu:diva-186337 (2022). Master student thesis.

48. Badža, M. M. & Marko, C?. B. Classification of brain tumors from MRI images using a convolutional neural network. Appl. Sci. 10, 1999 (2020).

49. Almalki, Y. E. et al. Isolated Convolutional-Neural-Network-Based Deep-Feature Extraction for Brain Tumor Classification Using Shallow Classifier. Diagnostics 12, 1793 (2022).

50. Sultan, H. H., Salem, N. M. & Al-Atabany, W. Multi-classification of brain tumor images using deep neural network. IEEE access 7, 69215–69225 (2019).

51. Singh, A., Sengupta, S. & Lakshminarayanan, V. Explainable deep learning models in medical image analysis. J. Imaging 6, 52 (2020).

